# Mapping of yaws endemicity in Ghana; Lessons to strengthen the planning and implementation of yaws eradication

**DOI:** 10.1101/2020.02.20.20025122

**Authors:** Laud Anthony Wihibeturo Basing, Moses Djan, Shirley Victoria Simpson, Yaw Adu-Sarkodie

**Affiliations:** Department of Clinical Microbiology, KNUST, Kumasi, Ghana; Health Information Unit, Municipal Health Directorate, Asamankese, West Akim, Ghana; Bacteriology Department, Noguchi Memorial Institute for Medical Research, Accra, Ghana

**Keywords:** Yaws, DPP, *Haemophilus ducrey*, Distribution, Ghana, *Treponema pallidum subsp pertenue*

## Abstract

**Introduction:** Yaws caused by *Treponema pallidum subsp pertenue* is a disease of poverty and affects communities where basic socio-economic amenities are lacking. With results showing that single dose azithromycin is effective in the treatment of yaws, the World Health Organisation introduced the Morges strategy with the intent to eradicate yaws by 2020. Ghana is one of the countries with the most yaws cases globally, and the National Yaws Eradication Program in Ghana intends to conduct Mass Drug Administration (MDA) of endemic communities in line with the Total Community Treatment plan of the Morges strategy. It is therefore important to map out endemic communities to ensure that MDA is both effective and financially efficient.

**Methods:** Children with suspected yaws lesions were actively selected from the recruitment sites (schools and communities). A full medical history, study site information including GPS coordinates, demographic data including communities of residence and clinical assessment were taken. Each of the clinically diagnosed children were screened using the DPP^®^ Syphilis Screen & Confirm Assay (DPP). Samples for PCR were collected by swabbing ulcerative lesions of participants and tested for *Treponema pallidum subsp pertenue* and *Haemophilus ducreyi DNA*

**Results:** In all, 625 children with a median age of 10 years were recruited into the study. While 401(64.2%) were DPP positive, only 141 of them had *Treponema pallidum subsp pertenue* DNA (TPE_DNA) accounting for 22.6% of those who were clinically diagnosed. Based on the DPP results, yaws was endemic in all the 4 study sites with participants from 88 communities in 13 districts in 4 regions in Ghana. There was no statistically significant difference between the various districts in terms of DPP results (x^2^=0.9364, p= 0.817) and 154 (24.6%) of those clinically diagnosed as yaws were positive for *Haemophilus ducreyi i* DNA.

**Conclusion:** Our study shows that communities endemic for yaws are also endemic for *Haemophilus ducreyi i*. Most yaws endemic communities were found at the border of other districts and regions. It is recommended that MDA should not only target endemic communities, it should target entire endemic districts as well as neighbouring districts in order to be effective.

## BACKGROUND

Yaws which is caused by *Treponema pallidum subsp pertenue* is a disease of poverty and affects communities where basic socio-economic amenities are lacking (1,2). Yaws is currently endemic in 13 countries although accurate data from these 13 countries as well as the 73 countries that were previously endemic are unavailable (3,4). An attempt by the World Health Organisation (WHO) to eradicate yaws led to a reduction of the global prevalence of yaws by 95% (5). However logistical challenges of injectable benzathine penicillin as well as weak primary health care systems resulted in the inability to get rid of the remaining 5% which resurged in several countries in the late 1970s (6).

Currently, 84% of yaws cases are concentrated in just three countries—Ghana, Papua New Guinea and the Solomon Islands and even in these countries’ yaws surveillance programmes report cases based on clinical diagnosis without confirmation via serology or PCR(7). Studies in endemic countries show that *Haemophilus ducreyi* and not *Treponema pallidum subsp pertenue* is responsible for many of the ulcers that had previously been reported as yaws (8–10). Diagnosis of yaws using the Dual Path Pathway (DPP), a serological method that detects both treponemal and non-treponemal antibodies is recommended and is currently in use in Ghana and other endemic countries (11–14).

With the results of a study in Papua New Guinea in 2012, showing that a single-dose of oral azithromycin is effective in treating yaws (15), the World Health Organization launched a roadmap that targets the eradication of yaws by 2020 (16). This global Strategy called the Morges Strategy has as one of its policies, a total community treatment (TCT) which implies the treatment of an entire endemic community, irrespective of the number of active clinical cases in those communities (17). However, it is important that this strategy is implemented in the context of the situation on the ground to ensure that Mass Drug Administration (MDA) is both effective and financially efficient. A previous MDA in the West Akim district showed a reduction of cases from 10.9% to 2.2% (18). However, a subsequent survey in the same district found pockets of infection all over the district(19). Proximity between communities and even districts as well as active interactions between people living in these communities and districts raises concern about whether MDA strategies should just target endemic communities or perhaps endemic districts and their immediate neighbours. It is therefore important to establish the extent and geographical distribution of yaws in districts where yaws is suspected to enable the national Yaws eradication Programme in Ghana to focus interventions on communities where transmission continues. Confirmation of the eradication of yaws will be difficult without such a mapping exercise in formerly endemic districts since, without this, unidentified pockets of infections will probably persist. Determining areas of yaws endemicity and mapping out these areas would essentially ensure prudent use of limited resources during mass drug administration processes. It would also provide baseline data on the determinants of spatial distribution of yaws in the study districts.

This study was therefore aimed at mapping up the distribution of yaws in the study districts as well as explore the determinants of the spatial distribution of yaws in order to facilitate completion of baseline mapping.

## METHODS

### Ethical Clearance

The study protocol was submitted for approval by the ethical review committee of the Ghana Health Service as well as institutional ethical bodies of London School of Hygiene and Tropical Medicine, Kwame Nkrumah University of Science and Technology, Noguchi Memorial Institute for Medical Research, the United States Centre for Disease Control and Prevention and the World Health Organization’s Ethical Review Committee. Only individuals who had lesions consistent with yaws who had consented and/or assented to participate in the trial were enrolled in the study. Participants and/or their parents or guardians were allowed to withdraw assent/consent at any time, without specified reason(s). Their rights to benefit from treatment however remained unchanged as a result of their withdrawal from the study.

### Study Communities

The study was conducted in communities in four yaws endemic districts in Ghana as well as communities in neighbouring districts that actively interacted with these endemic communities. Figure 1, shows the map of Ghana, highlighting the study districts.

**Fig 1:**
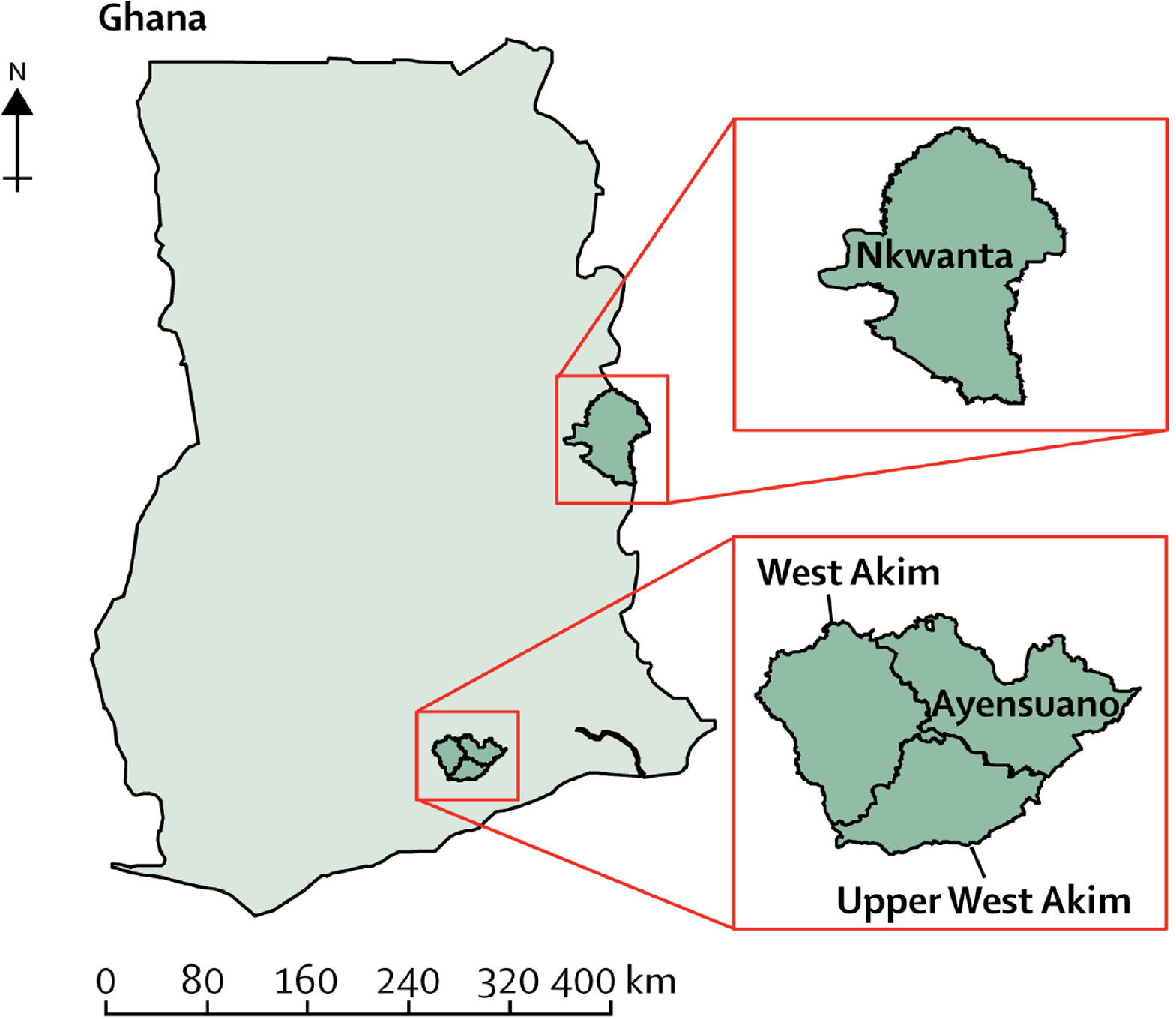
A map of the study districts and their location in Ghana (19)

The Ayensuanor district with a population of 77,193 lies within Latitudes 5°45’N and 6°5’N and Longitudes 0°15’W and 0° 45’W. It is in the southern part of the Eastern Region and shares boundaries with Suhum Municipality to the North; Nsawam Adoagyiri Municipality to the South; Akwapem South District to the East and Upper West Akim District and West Akim Municipality to the West. The district is made up of 9 sub districts namely; Asuboi, Manase, Coaltar, Marfo, Anum Apapam, Kofi Pare, Kuano, Dokrochiwa and Teacher Mante.

The West Akim Municipal Assembly lies between longitudes 0°25’ West and 0°47’ West and 60° 0’ North. The municipality shares boundaries with Denkyembour District Assembly and East Akim Municipal Assembly to the North, Birim Central Municipal Assembly to the West and Ayensuoano and Upper West Akim District Assemblies to the East. It has a population of about 108,298

The Upper West Akim District lies between longitudes 0°25’ West and 0°47’ West and latitudes 5°40’ North and 6°00’ North. It shares boundaries with Ayensuano District to the east, West Akim Municipality to the north, Nsawam Adoagyiri Municipality to the southeastern part, Ga South Municipality to the south and Awutu-Afutu Senya District in the Central region to the west. This district also shares boundaries with the Central and Greater Accra Regions. The district with a population of 87,051 is made up of 7 sub districts, namely Abamkrom, Adeiso, Asuokaw, Mepom, Nyanoa, Odumkyere Darmang and Okurase

The Nkwanta North District being one of the twenty-five (25) districts in the Volta Region, is located between Latitude 7°30’N and 8°45’N and Longitude 0°10’W and 045’E. The district shares boundaries with the Nanumba South District to the North, Republic of Togo to the East, Kpandai District to the West and Nkwanta South District to the South.

### Definition of yaws endemicity

A community was defined as endemic for yaws if one or more persons tested positive for the DPP test. The DPP test is an alternative to the traditional RPR and TPHA tests for the diagnoses of Treponemal infections. The TPHA is highly specific but frequently remains positive for life following infection, regardless of treatment or natural clearance. The RPR, on the other hand, is less specific but reflect active disease more accurately, although positive RPR results may also be seen in serofast patients. The DPP test is based on a combination of both tests with the T1 line being that for the Treponemal line akin to the TPHA test and the T2 line being the non-Treponemal line akin to the RPR and a sample to answer time of 15 minutes.

### Sample and Data collection

Patients (both genders) with clinically diagnosed yaws lesions were actively selected from the recruitment sites (schools and communities). A full medical history, study site information including GPS coordinates, demographic data including communities of residence and clinical assessment, were performed and all information recorded onto a standardized Patient Record Form.

Each of the clinically diagnosed children were screened using the DPP^®^ Syphilis Screen & Confirm Assay (Chembio Diagnostic Systems Inc. Medford, NY) as described by Causer et al (12), Samples for PCR were collected by swabbing ulcerative lesions and scrapping from papilloma of DPP positive participants to confirm *Treponema pallidum subsp pertenue*. Dacron-tipped cotton wool swabs pressed and rolled over the ulcerated lesions whilst disposable plastic curettes were used to collect samples from papillomatous lesions. The samples were carefully suspended in Assay Assure transport medium (Thermo Fisher Scientific, Waltham, MA) and stored between 4-8°C until it was transported to Noguchi Memorial Institute for Medical Research. Samples for diagnostic confirmation were handled according to existing procedures recommended by the World Health Organization. All samples in this study were analyzed at the Centers for Disease Control and Prevention, Atlanta, USA

### Laboratory analysis

DNA from the swab samples was extracted using the manufacturer’s instructions. The molecular differentiation of *Treponema pallidum subsp pertenue* from *Treponema pallidum subsp pallidum* and *Treponema pallidum subsp endemicum* was achieved using a real-time Triplex PCR containing primers and TaqMan probes targeting the tp0858 gene for *Treponema pallidum subsp pertenue*, which encodes a hypothetical protein and two areas of the tprI (tp0620) gene for *Treponema pallidum subsp endemicum Treponema pallidum subsp pallidum* as described by Chi et al (20)

The triplex PCR to detect *Mycobacterium ulcerans, Haemophilus ducreyi* and RNA was performed with a 10-mL sample of DNA in a 25-mL reaction volume containing 12.5-mL of PerfeCTa Multiplex qPCR SuperMix (Quanta Biosciences, Gaithersburg, MD) and a volume of 0.2ul of each primers and Taqman hybridization probe with a florescent dye (calRed for M. ulcerans, HEX for Haemophilus ducreyi i and Quas670 for RNP-Qiagen, USA).

### Data Analysis

All the data obtained from the districts were recorded into Microsoft® Excel and analysis was performed using the Stata 14 statistical software (StataCorp. 2015. Stata Statistical Software: Release 14. College Station, TX: StataCorp LP) after adding laboratory data. Demographic Characteristics and their association with *Treponema pallidum subsp pertenue* and *haemophilus ducreyi* infection, were analyzed using the Fischer’s exact test or Chi square test where necessary. Continuous variables were expressed as medians with their inter-quartile ranges (IQR). Using logistic regression, bivariate analysis was performed to estimate the odds ratios (OR) and their 95% CI. To determine the independent risk factors of *Treponema pallidum subsp pertenue* infections, socio-demographic variables that were significant at probability value (p) < 0.1 from the bivariate analysis were entered into an unconditional multiple logistic regression model. A forward and backward stepwise approach was used for selection of significant variables while adjusting for all variables in the model. All variables from the model were expressed as the adjusted odd ratios (OR) and 95% confidence interval (CI). A two-sided p-value of less than 0.05 was considered significant. Spatial analysis and mapping were done using ArcGIS *10*.*2*.*2* (ESRI 2011. ArcGIS Desktop: Release 10. Redlands, CA: Environmental Systems Research Institute). GPS coordinates were extracted and saved in a shape file which was analysed and mapped.

## RESULTS

### Demography

In all, 625 children with a median age of 10 years were recruited into the study. 70.9% of these children were males whilst 29.1% were females. These children were recruited in schools and communities based on clinical yaws like lesions. 82.4% of the lesions were ulcers whilst 17.6% were papillomata. The leg was the part of the body with the highest number of legions (81.9%), whilst the arm, the back and the neck or face were other areas that had lesions. The trend was similar across the various districts. Out of the 146 recruited from Ayensuanor district, accounting for 22.4% of the children recruited in Ghana, 71.2% were males, as against 28.8% females. The median age of those recruited in Ayensuanor was 10, with ulcers accounting for 78.8% of the lesions found in the district. The leg was the part of the body with the highest number of lesions (77.4%). This trend is similar across all the districts, except for the median age in Nkwanta North which was 9 years as against a median age of 10 across the various districts. The Upper West Akim District which recruited 31.8% of the total number of children recruited in the Ghana study hence making it the district with the highest number of enrolled children in the study had more children with legions at their back than with lesions on their face or necks. For each of the districts, the leg had more lesions, followed by the arm, then the face or neck and then their backs, however in the Upper West Akim District, the more children had lesions on their backs than on their faces.

### Treponemal and Non-Treponemal tests in children with yaws lesions

Out of the 625 children who were clinically diagnosed with yaws, 401(64.2%) were DPP positive. Only 141 of them had *Treponema pallidum subsp pertenue* DNA (TPE_DNA) accounting for 22.6% of those who were clinically diagnosed. 154 (24.6%) of those clinically diagnosed as yaws were positive for *Haemophilus ducreyi* DNA (H. ducreyi_DNA). In Ayensuanor district, 93 of the 146 (63.7%) children clinically diagnosed with yaws were positive using the DPP test as against 92 (65.2%) in Nkwanta North, 138 (69.3%) in Upper West Akim District and 78 (56,1%) in the West Akim District. There was no statistically significant difference between the various districts in terms of DPP results (x^2^=0.9364, p= 0.817). Sixty (60) of the children clinically diagnosed with yaws in Ayensuanor tested positive for TPE_DNA giving a prevalence of 41.1%. In the Upper West Akim district with had the highest number of clinically diagnosed yaws, 45 (22.6%) tested positive for TPE_DNA while Nkwanta North and West Akim District both had 18 children testing positive accounting for 12.8% and 12.9% respectively of the children clinically diagnosed with yaws. A chi^2^ test to detect if there was a significant difference between TPE_DNA results across the various districts showed that there was a significant difference in TPE_DNA positive cases across the various districts (x^2^=30.6777, p= <0.001). On the other hand, 47 (33.8%) of clinically diagnosed yaws infections tested positive for *Haemophilus ducreyi* in the West Akim district, 52 (26.1%) in the Upper West Akim District, 30 (21.9%) in the Ayensuanor District and 18 (12.8%) in the Nkwanta North District. There was no statistically significant difference in *H. ducreyi_*DNA results across the various districts (x^2^=5.6446, p= 0.130) (**Table 1**)

**Table 1:**
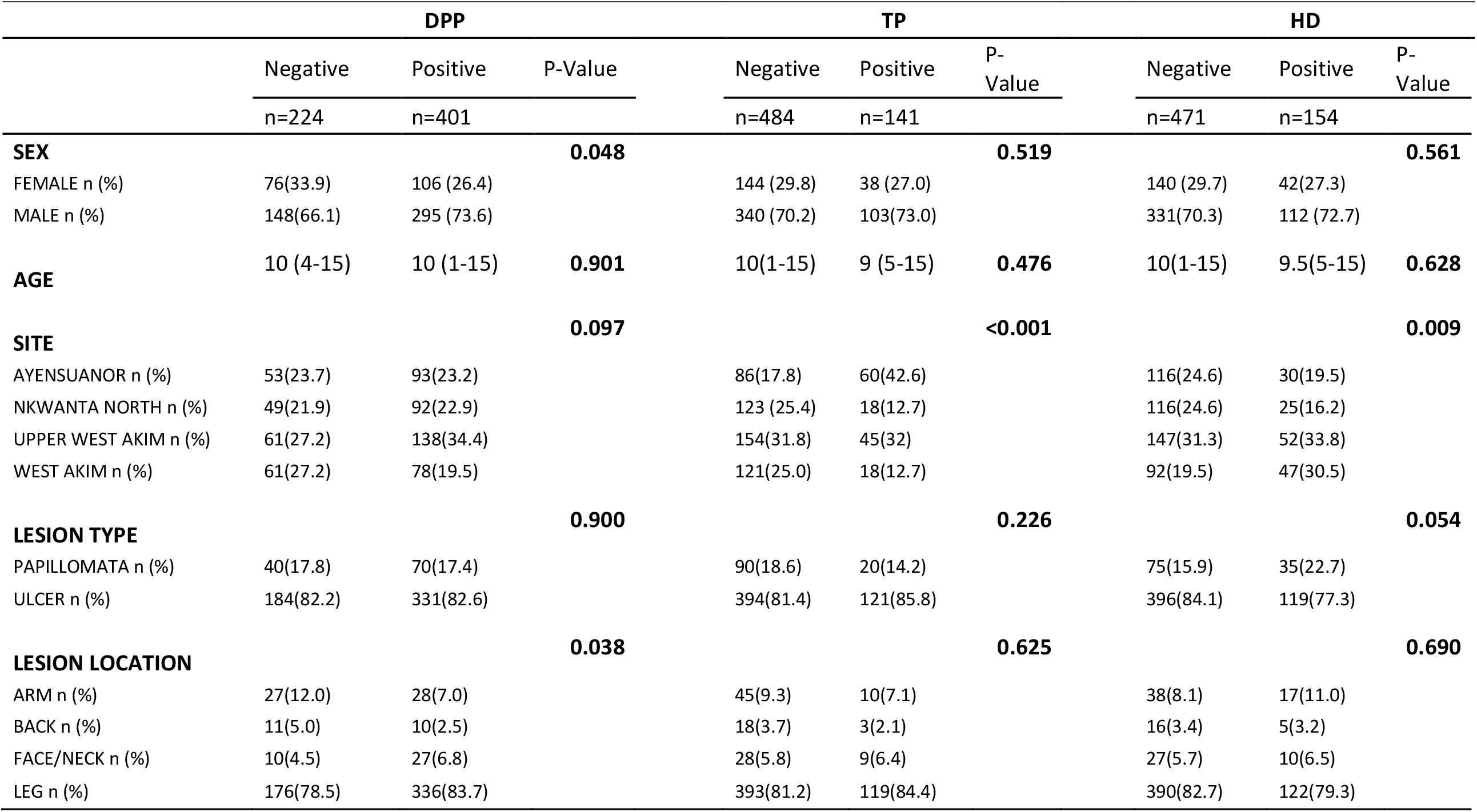
Demographic characteristics associated with DPP, TP and HD testing

|  | DPP |  |  | TP |  |  | HD |  |  |
| --- | --- | --- | --- | --- | --- | --- | --- | --- | --- |
|  | Negative | Positive | P-Value | Negative | Positive | P-Value | Negative | Positive | P-Value |
|  | n=224 | n=401 |  | n=484 | n=141 |  | n=471 | n=154 |  |
| **SEX** |  |  | **0.048** |  |  | **0.519** |  |  | **0.561** |
| FEMALE n (%) | 76(33.9) | 106 (26.4) |  | 144 (29.8) | 38 (27.0) |  | 140 (29.7) | 42(27.3) |  |
| MALE n (%) | 148(66.1) | 295 (73.6) |  | 340 (70.2) | 103(73.0) |  | 331(70.3) | 112 (72.7) |  |
| **AGE** | 10 (4-15) | 10 (1-15) | **0.901** | 10(1-15) | 9 (5-15) | **0.476** | 10(1-15) | 9.5(5-15) | **0.628** |
| **SITE** |  |  | **0.097** |  |  | **<0.001** |  |  | **0.009** |
| AYENSUANOR n (%) | 53(23.7) | 93(23.2) |  | 86(17.8) | 60(42.6) |  | 116(24.6) | 30(19.5) |  |
| NKWANTA NORTH n (%) | 49(21.9) | 92(22.9) |  | 123 (25.4) | 18(12.7) |  | 116(24.6) | 25(16.2) |  |
| UPPER WEST AKIM n (%) | 61(27.2) | 138(34.4) |  | 154(31.8) | 45(32) |  | 147(31.3) | 52(33.8) |  |
| WEST AKIM n (%) | 61(27.2) | 78(19.5) |  | 121(25.0) | 18(12.7) |  | 92(19.5) | 47(30.5) |  |
| **LESION TYPE** |  |  | **0.900** |  |  | **0.226** |  |  | **0.054** |
| PAPILLOMATA n (%) | 40(17.8) | 70(17.4) |  | 90(18.6) | 20(14.2) |  | 75(15.9) | 35(22.7) |  |
| ULCER n (%) | 184(82.2) | 331(82.6) |  | 394(81.4) | 121(85.8) |  | 396(84.1) | 119(77.3) |  |
| **LESION LOCATION** |  |  | **0.038** |  |  | **0.625** |  |  | **0.690** |
| ARM n (%) | 27(12.0) | 28(7.0) |  | 45(9.3) | 10(7.1) |  | 38(8.1) | 17(11.0) |  |
| BACK n (%) | 11(5.0) | 10(2.5) |  | 18(3.7) | 3(2.1) |  | 16(3.4) | 5(3.2) |  |
| FACE/NECK n (%) | 10(4.5) | 27(6.8) |  | 28(5.8) | 9(6.4) |  | 27(5.7) | 10(6.5) |  |
| LEG n (%) | 176(78.5) | 336(83.7) |  | 393(81.2) | 119(84.4) |  | 390(82.7) | 122(79.3) |  |

### Factors associated with *Treponema pallidum subsp pertenue*

To determine the factors that could best predict exposure to *Treponema pallidum subsp pertenue* infection, a logistic regression model allowing for adjustment of all factors associated with *Treponema pallidum subsp pertenue* infection at p-values <0.05 were performed. The values selected were staying in Ayensuanor, West Akim, Upper West Akim District, DPP, HD lesions located at the back were all identified as the independent risk factors of Treponema pallidum pertenue (Table 2).

**Table 2:** Factors associated with Treponema pallidum pertenue infection

|  | Positive<br>n (%) | Negative<br>n (%) | Crude<br>OR (95% CI) | Adjusted<br>OR (95% CI) | P<br>Value |
| --- | --- | --- | --- | --- | --- |
| **Age** |  |  |  |  |  |
| <5 n (%) | 0(0%) | 2 (0.41%) | 0.000 |  |  |
| 6 to 8 n (%) | 55 (39.01)) | 146 (30.17) | 1.000 |  |  |
| 9 to 12 n (%) | 66 (46.81) | 245 (50.62) | 1.04 (.68-1.59) | 1.09 (0.70-1.68) | 0.87 |
| 13 to 15 n (%) | 20 (14.18) | 91 (18.80) | 1.15 (0.67-1.98) | 1.24 (0.71-2.16) | 0.624 |
| **Site** |  |  |  |  |  |
| Nkwanta n (%) | 24 (17.02) | 117 (24.17) | 1.000 |  |  |
| Ayensuanor n (%) | 49 (34.75) | 97 (20.04) | 9.95 (4.69-21.09) | 10.60 (4.97-22.60) | <0.001 |
| West Akim n (%) | 40 (28.37) | 99 (20.45) | 3.02 (0.03-0.13) | 3.09 (1.43-6.67) | 0.004 |
| UWA n (%) | 28 (19.86) | 171 (35.33) | 5.72 (2.65-12.35) | 5.94 (2.74-12.87) | <0.001 |
| **Sex** |  |  |  |  |  |
| Female n (%) | 28 (19.86) | 154 (31.82) | 1.000 |  |  |
| Male n (%) | 113 (80.14) | 330 (68.18) | 1.15 (0.75-1.75) | 1.27 (0.77-1.82) | 0.683 |
| **DPP** |  |  |  |  |  |
| Negative n (%) | 318 (65.70) | 166 (34.30) | 1.000 |  |  |
| Positive n (%) | 83 (58.87) | 58 (41.13) | 58.89 (14.42-240.54) | 63.61 (15.49-261.14) | <0.001 |
| **HD** |  |  |  |  |  |
| Negative n (%) | 132 (27.27) | 352 (72.73) | 1.000 |  |  |
| Positive n (%) | 22 (15.60) | 119 (84.40) | 0.18 (.092-.35) | 0.19 (0.096-0.37) | <0.001 |
| **Lesion type** |  |  |  |  |  |
| Papilomata n (%) | 25 (17.73) | 85 (17.56) | 1.000 |  |  |
| Ulcers n (%) | 116 (82.27) | 399 (82.44) | 1.20 (0.72-2.00) | 1.23 (0.86-2.29) | 0.362 |
| **Lesion Location** |  |  |  |  |  |
| Arm n (%) | 11 (7.80) | 44 (9.09) | 1.000 |  |  |
| Face/Neck n (%) | 2 (1.42) | 19 (3.93) | 1.45 (0.52-3.99) | 1.69 (0.59-4.83) | 0.478 |
| Back n (%) | 8 (5.67) | 29 (5.99) | 3.38 (1.12-10.17) | 4.31 (1.38-13.45) | 0.027 |
| Leg n (%) | 120 (85.11) | 392 (80.99) | 1.27 (0.62-2.61) | 1.35 (0.65-2.82) | 0.507 |

### Mapping yaws endemicity in the study communities

Yaws was endemic in all the 4 study sites with participants from 86 communities in 13 districts in 4 regions in Ghana. In the Nkwanta North District, patients surveyed came from both the Nkwanta North District and the Nkwanta South District. A total of 10 communities were surveyed with participants being DPP positive in all 10 communities surveyed. The communities with the most DPP positive cases were from Sibi Central, Sibi Hilltop and Lamina in the Nkwanta South District and Krachi Akura in the Nkwanta North District. Participants from the Upper West Akim District came from 30 communities with DPP positive participants coming from 29 of those communities from 5 districts in 3 regions. Communities with high DPP positive cases include Brofuyedu, Obinimda, Esusu, Okurase and Siwunkyemu. Most of the DPP positive cases from the West Akim Municipality came from participants who resided outside the municipality. In all DPP positive participants were from 28 out of the 29 communities surveyed in 4 districts. While communities like Fawomanye and Otwikrom in the municipality had high number of DPP positive cases, most of the DPP positive cases came from communities like Agona Kwasikum in the Central Region of Ghana. In the Ayesuanor District, Achiansah, Apau Wawase and its surrounding communities and Teacher Mante were all positive for DPP. However, a few cases came from Alaafia in the Upper West Akim District.

All the TPE_DNA positive cases were from communities with DPP positive cases. In the Nkwanta North District, 4 communities out of the 10 DPP positive communities were positive for TPE_DNA. In the Upper West Akim District, 10 out of 29 DPP positive communities were positive for TPE_DNA. In Ayensuanor, TPE_DNA positive cases were concentrated in 4 communities out of the 19 yaws endemic communities and in the West Akim district, 13 out of 28 DPP positive communities were positive for TPE_DNA.

*Haemophilus ducreyi* DNA positive cases were also endemic in communities which had DPP positive cases. Lamina in the Nkwanta North District recorded the highest number of DPP positive cases as well as the highest number of *Haemophilus ducreyi* cases. In the Upper West Akim district, the distribution of *Haemophilus ducreyi* was all over communities which had cases of DPP positive lesions except in Esikasu Odumase which recorded three *Haemophilus ducreyi* positive cases although it did not have any DPP positive case. In the West Akim District, Fawomanye which recorded the highest number of DPP cases also recorded the highest number of *Haemophilus ducreyi* cases while in Ayensuanor, most of the communities that had DPP positive cases also had *Haemophilus ducreyi* positive cases except for Kuano which two cases testing positive for *Haemophilus ducreyi* although there were no DPP positive cases recorded in the community. In all, 4, 19, 17 and 7 communities in the Nkwanta North, Upper West Akim, West Akim and Ayensuanor district respectively had *Haemophilus ducreyi* positive cases out of the 10, 3o, 29 and 19 communities surveyed.

## Discussion

In our study, there were more males in all the districts than females accounting for 70.9% of the cases. The median age of the children was 10 on the average and 10 in all the districts except Nkwanta North which had a median age of 9. Over 80% of the lesions were ulcers found on the legs. Yaws cases appear to affect mostly children between 2 and 15 years old, who are also considered as the reservoir for infections(21). It appears that rather more males than females suffer from the yaws. This because boys are more active than girls and therefore suffer more traumas while playing with each other and hence can transmit the infections faster. It is considered that transmission occurs through direct skin contact with a fluid from an infected lesion. In addition to this, there usually are extensive areas of vegetation (bush) in these rural communities, which may increase the chances of injury to legs and feet and, therefore, liability of infection. The mainstay of these communities is farming and since children help their parents on the farms, personal injuries on the farm could help spread the infection

As seen in Table 1, 64.2 percent of those who were clinically diagnosed with yaws were DPP positive. There was no difference in DPP results across the various districts. Our study had quite a high agreement between clinical diagnosis and DPP testing (64.2%). This is quite different from studies comparing clinical diagnosis and serological results. Marks et al. (2014) had an agreement of 29.6% in the Solomon Islands. Ayove et al. (13) in Papua New Guinea had an agreement of 41% between serological methods and clinical diagnosis. Clinical diagnosis of yaws although very subjective (13) is largely dependent on the actual prevalence of the yaws in the community as well as the experience of the person making the clinical diagnosis. Kwakye-Maclean et al. (22) in Ghana, had as high as 87.6% seroprevalence in the West Akim District (Upper West Akim and West Akim) before Mass Drug Administration to these patients and their contacts. Communities in the communities enrolled in the study are endemic for yaws and have been for several years, most of those who make clinical diagnosis have been trained to help in surveillance activities and have been doing that for many years, combined with visual aids for yaws, it is expected that clinical diagnosis would be quite consistent with serological diagnosis.

In our study, 22.6% of those clinically diagnosed with yaws were positive for TPE_DNA. This result is consistent with Mitja et al.(23) in Papua New Guinea with TPE_DNA in 21.1% of the cases. The results, however, is higher than Chi et al. (20) who found TPE_DNA in 15.5% in Vanuatu, and Houinei et al. (24) who found TPE_DNA in only 1% of the cases in Papua New Guinea, However, this study was done by swabbing the skin of asymptomatic children, without ulcers or papillomata. The result in this study was, however, lower than the 37% found in a community-based cohort study in Lihir Island, Papua New Guinea, from October 2014 through May 2016 (25).

There was a significant difference in TPE_DNA positive cases across the various districts (x^2^=30.6777, p= <0.001). Whereas 41.1% of the cases in Ayensuanor District were positive for TPE_DNA, only 12.8% of children in Nkwanta North were positive for TPE_DNA. Several factors could account for the inter-country and inter-district variation in TPE_DNA positive cases. Grange et al. (26) recommend that to have effective detection of treponemal DNA from swab samples, a sterile unmoistened dacron swab on the lesion over a 5-by 5-cm area should be taken. However, with a multicentre trial and with different people taking swab samples, the likelihood of variation is increased. The Ayensuanor district is unique because it borders both the West Akim District and the Upper West Akim district. While these two districts were known to be highly endemic for the Yaws and had gone through an MDA. Ayensuanor has never gone through an MDA, and it is therefore not surprising that they have a high number of PCR positives. Upper West Akim district is quite impressive because although they went through the MDA, 22.6% still had TPE_DNA.

In our study, *Haemophilus ducreyi* was found to be positive in 24.6% of the cases, with 21.9% being found in Ayensuanor, 17.7% being found in Nkwanta North, 26.1% being found in Upper West Akim and 33.8% being found the West Akim District. There was no statistically significant difference in the results across the various districts (x^2^=0.9364, p= 0.817). The results in the study were similar to results obtained in Papua New Guinea by Houinei et al. (24) (21%) and by Marks et al. (9) in the Solomon Islands (32%). The result in this study however differed from results by Mitja et al (23) and Gonzales-Bieras et al (25) in Papua New Guinea (46.7% and 53% respectively) and one study conducted in the Northern Part of Ghana (10) which had H ducreyi in 9 lesions out of 98 lesions. It is instructive to note that no TPE_DNA was found in that particular study. It is also instructive to note that in our study, aside Ayensuanor which had more TPE_DNA positive cases than *Haemophilus ducreyi* positive cases, all the districts had more *Haemophilus ducreyi* cases than TPE_DNA positive cases.

In our study, we expected that the chronic ulcers in children younger than 15 years would be mainly caused by T pallidum pertenue or even if there were some caused by *Haemophilus ducreyi*, it was expected that those caused by T pallidum pertenue would be greater than those caused by *Haemophilus ducreyi*. However, our molecular assessment of ulcer cause showed that *Haemophilus ducreyi* is a very common trigger of chronic skin ulcer and in our study and the assessment of other studies held in other regions in the world (Papua New Guinea and the Solomon Islands) show that *Haemophilus ducreyi i* is endemic in yaws endemic communities (8,25,27).

The Morges strategy for the eradication of yaws by 2020 is to give one round of total community treatment with azithromycin to endemic communities, followed by regular surveys to identify and treat new cases and their contacts (28). To effectively do this, it is important to know exactly where yaws can be found, and which communities are susceptible. In our study, yaws was found in thirteen (13) districts in Four (4) regions in Ghana instead of the study sites of 4 districts in 2 regions. This is because there were patients from communities outside the study areas who interact daily with the four endemic districts, which shows that in the eradication of yaws, surrounding communities must also be targeted if the fight against yaws is to be won. Most of the endemic communities are found at the boundaries with other districts or even regions. As can be seen in Fig 2c and Fig 3b, some communities are actually found between two districts and as such interact with people from the other nearby districts on regular basis. Upper West Akim district had the highest number of cases recruited into the study accounting for 31.8% of the study. A previous study conducted a mass drug administration(MDA) in the then West Akim District (Comprising the current Upper West Akim District and the West Akim District) (22), yet these two districts combined accounted for more than half of the cases in this study as compared to Ayensuanor and Nkwanta North which has never gone through Mass Drug Administration. This shows that Upper West Akim and West Akim continue to be showcases of yaws like lesions even after the MDA. In some of the communities, children live in one district and go to school or to the farm in another district. For example, Brofoyedru is in the Upper West Akim District but it shares a boundary with the Agona West District of the central Region. A lot of the inhabitants have farms that are in the central region and so children when they are not in school, spend a lot of time with their families in the Central region. Treating children in Brofoyedru without treating children in neighbouring communities in the Agona West District would therefore be quite ineffective and may account for the reason the Upper West Akim and the West Akim districts, beneficiaries of MDAs still had very high yaws prevalence.

**Fig: 2:**
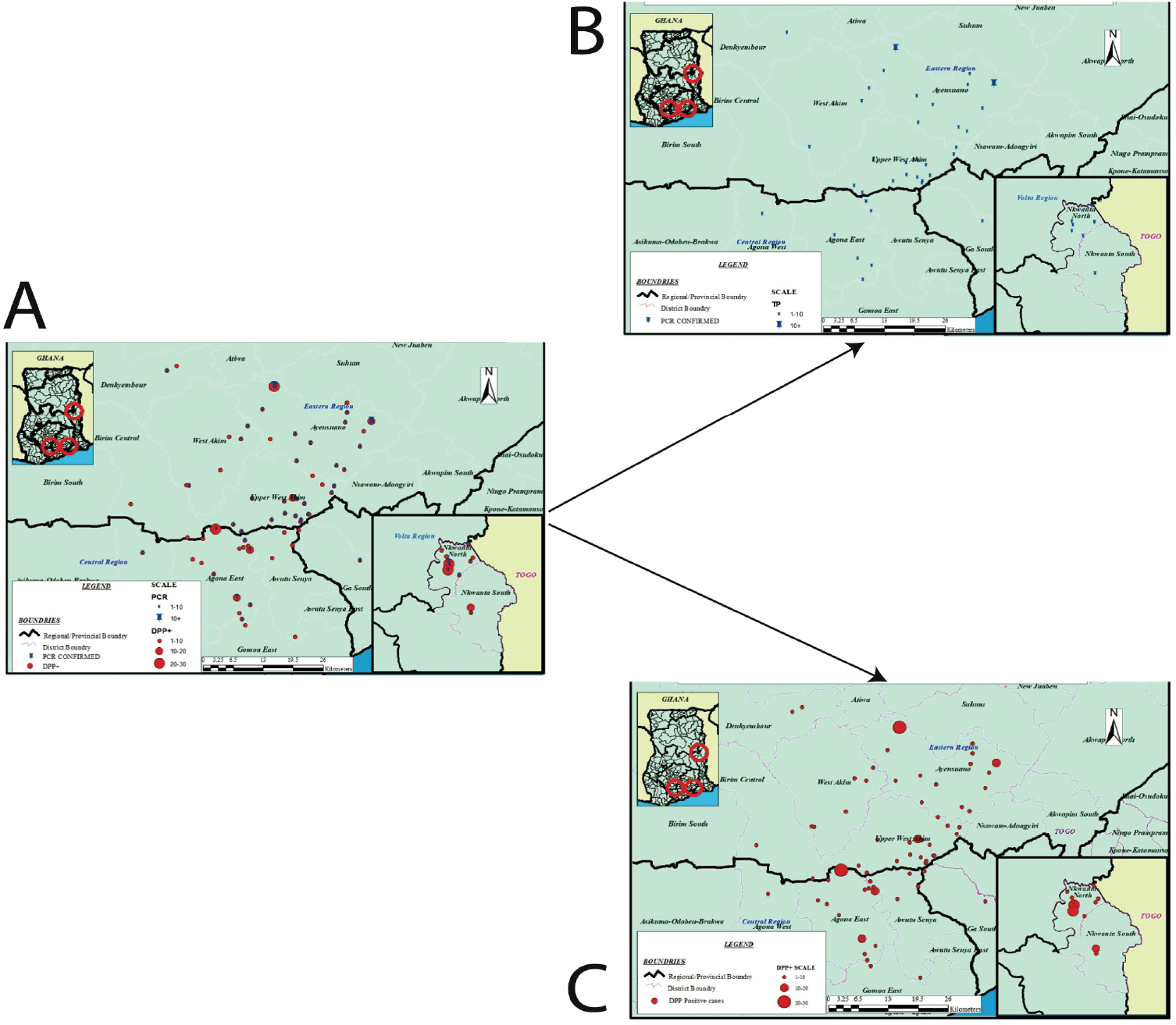
Geographical distribution of *Treponema pallidum subsp pertenue* **A:** A map showing the distribution of DPP positive cases and TPE_DNA cases. **B**: A map showing the distribution of TPE_DNA cases only **C**: A map showing the distribution of DPP positive cases only

**Fig: 3:**
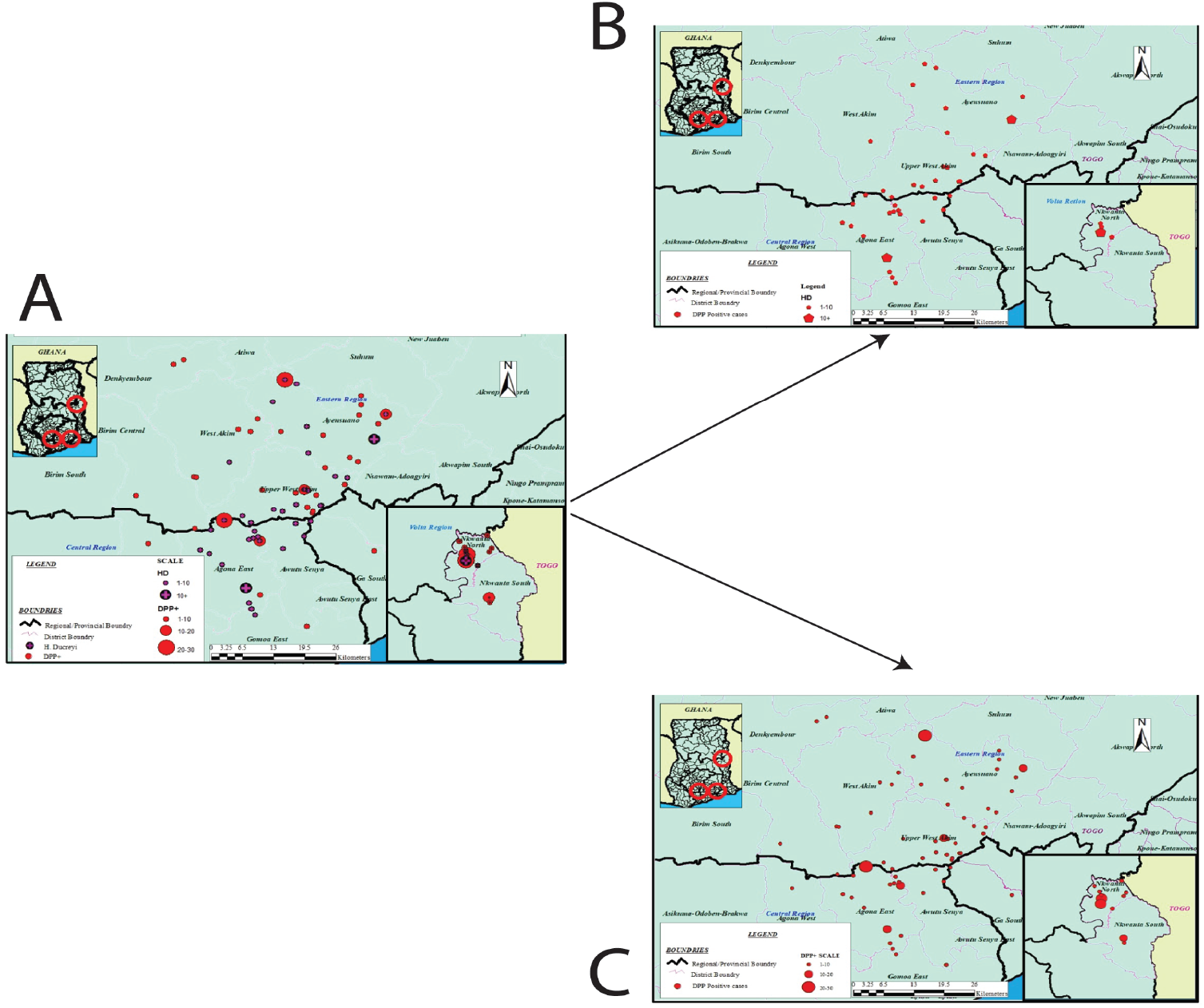
Geographical distribution of *Haemophilus ducreyi* **A:** A map showing the distribution of *H. ducreyi*_DNA and DPP positive cases. **B**: A map showing the distribution of *H. ducreyi*_DNA cases only **C**: A map showing the distribution of DPP positive cases only

Our study in Fig, 2 and 3 shows the geographical distribution of *Treponema pallidum subsp pertenue* and *Haemophilus ducreyi* infection in the communities. What is interesting to note is that communities that are endemic for yaws are also endemic for *Haemophilus ducreyi*. Communities like Lamina and Sibi in Nkwanta North, Okurase, Siwunkyemu, Esusu, Brofu Yedru, Akokwa etc in the Upper West Akim District were endemic for both infections. In the West Akim District, Fawomanye and Otwekrom while Achiansah, Appau Wawase and Teacher Mante in the Ayensuanor district had both infections. It is also noteworthy that several communities which tested DPP positive but were negative for TPE_DNA were also H. ducreyi_DNA negative. These include communities like Kwaboanta, Akokokrom, Esaase, Sasekrom, Breku Akura, Damako Papaye and kabre Akura. Communities that were either Yaws or Haemophilus ducreyi Positive but not both were found have neighboring communities that were the exact opposite of this. This included communities like Asuotewene and Ayensuako which had TPE-DNA positive and *Haemophilus ducreyi* positive respectively. With the above data, it is therefore important that any eradication effort targeted at *Treponema pallidum subsp pertenue* must also target *Haemophilus ducreyi*. Since studies have shown successful treatment of Haemophilus ducreyi with a single dose azithromycin (29), mass drug administration of azithromycin to children in these communities as well as in neighboring communities as well as contacts is essential in the eradication of yaws in Ghana.

## CONCLUSION

Yaws endemic communities were mapped out in our study with a number of patients coming from communities and Districts outside but close to the study sites. Mapping out the geographical distribution of yaws and *Haemophilus ducreyi* shows that communities endemic for yaws are also endemic for *Haemophilus ducreyi*. Most yaws endemic communities were found at the border of other districts and regions. It is recommended that MDA should not only target endemic communities, it should target entire endemic districts as well as neighbouring districts in order to be effective.

## Data Availability

All the data referred to in the manuscript are available and would be made available on request

## References

1. Capuano C, Ozaki M. Yaws in the Western pacific region: a review of the literature. J Trop Med. 2011 Dec 22;2011:642832.

2. Fitzpatrick C, Asiedu K, Jannin J. Where the road ends, yaws begins? The cost-effectiveness of eradication versus more roads. PLoS Negl Trop Dis. 2014; 8(9):e3165.

3. Kazadi WM, Asiedu KB, Agana N, Mitjà O. Epidemiology of yaws: an update. Clin Epidemiol. 2014;6:119–28.

4. Mitjà O, Marks M, Konan DJP, Ayelo G, Gonzalez-Beiras C, Boua B, et al. Global epidemiology of yaws: a systematic review. Lancet Glob Heal. 2015;3(6):e324–31.

5. Asiedu K, Fitzpatrick C, Jannin J. Eradication of Yaws: Historical Efforts and Achieving WHO’s 2020 Target. PLoS Negl Trop Dis. 2014;8(9).

6. Rinaldi A. Yaws eradication: facing old problems, raising new hopes. PLoS Negl Trop Dis. 2012;6(11):e1837.

7. Mabey D. Mapping the geographical distribution of yaws. Lancet Glob Heal. 2015; 3(6):e300–1.

8. Mitjà O, Lukehart SA, Pokowas G, Moses P, Kapa A, Godornes C, et al. Haemophilus ducreyi as a cause of skin ulcers in children from a yaws-endemic area of Papua New Guinea: a prospective cohort study. Lancet Glob Heal. 2014;2(4):e235–41.

9. Marks M, Chi K-H, Vahi V, Pillay A, Sokana O, Pavluck A, et al. Haemophilus ducreyi associated with skin ulcers among children, Solomon Islands. Emerg Infect Dis. 2014;20(10):1705–7.

10. Ghinai R, El-Duah P, Chi K-H, Pillay A, Solomon AW, Bailey RL, et al. A cross-sectional study of “yaws” in districts of Ghana which have previously undertaken azithromycin mass drug administration for trachoma control. Lammie PJ, editor. PLoS Negl Trop Dis. 2015;9(1):e0003496.

11. Yin Y-P, Chen X-S, Wei W-H, Gong K-L, Cao W-L, Yong G, et al. A dual point-of-care test shows good performance in simultaneously detecting nontreponemal and treponemal antibodies in patients with syphilis: a multisite evaluation study in China. Clin Infect Dis. 2013; 56(5):659–65.

12. Causer LM, Kaldor JM, Conway DP, Leslie DE, Denham I, Karapanagiotidis T, et al. An evaluation of a novel dual treponemal/nontreponemal point-of-care test for syphilis as a tool to distinguish active from past treated infection. Clin Infect Dis. 2015; 61(2):184–91.

13. Ayove T, Houniei W, Wangnapi R, Bieb S V., Kazadi W, Luke LN, et al. Sensitivity and specificity of a rapid point-of-care test for active yaws: A comparative study. Lancet Glob Heal. 2014;2(7):e415–21.

14. Marks M, Yin Y-P, Chen X-S, Castro A, Causer L, Guy R, et al. Metaanalysis of the Performance of a Combined Treponemal and Nontreponemal Rapid Diagnostic Test for Syphilis and Yaws. Clin Infect Dis. 2016; 63(5):627–33.

15. Mitjà O, Hays R, Ipai A, Penias M, Paru R, Fagaho D, et al. Single-dose azithromycin versus benzathine benzylpenicillin for treatment of yaws in children in Papua New Guinea: An open-label, non-inferiority, randomised trial. Lancet. 2012;379(9813):342–7.

16. Marks M. Yaws: towards the WHO eradication target. Trans R Soc Trop Med Hyg. 2016;110(6):319–20.

17. Marks M, Kwakye-Maclean C, Doherty R, Adwere P, Aziz Abdulai A, Duah F, et al. Knowledge, attitudes and practices towards yaws and yaws-like skin disease in Ghana. Huppert A, editor. PLoS Negl Trop Dis. 2017

18. Abdulai AA, Agana-Nsiire P, Biney F, Kwakye-Maclean C, Kyei-Faried S, Amponsa-Achiano K, et al. Community-based mass treatment with azithromycin for the elimination of yaws in Ghana—Results of a pilot study. Azman AS, editor. PLoS Negl Trop Dis. 2018;12(3):e0006303. Available from: https://dx.plos.org/10.1371/journal.pntd.0006303

19. Marks M, Mitjà O, Bottomley C, Kwakye C, Houinei W, Bauri M, et al. Comparative efficacy of low-dose versus standard-dose azithromycin for patients with yaws: a randomised non-inferiority trial in Ghana and Papua New Guinea. Lancet Glob Heal. 2018;6(4):e401–10.

20. Chi K-H, Danavall D, Taleo F, Pillay A, Ye T, Nachamkin E, et al. Molecular differentiation of Treponema pallidum subspecies in skin ulceration clinically suspected as yaws in Vanuatu using real-time multiplex PCR and serological methods. Am J Trop Med Hyg. 2015;92(1):134–8.

21. Marks M, Sokana O, Nachamkin E, Puiahi E, Kilua G, Pillay A, et al. Prevalence of Active and Latent Yaws in the Solomon Islands 18 Months after Azithromycin Mass Drug Administration for Trachoma. Vinetz JM, editor. PLoS Negl Trop Dis. 2016;10(8):e0004927.

22. Kwakye-Maclean C, Agana N, Gyapong J, Nortey P, Adu-Sarkodie Y, Aryee E, et al. A Single Dose Oral Azithromycin versus Intramuscular Benzathine Penicillin for the Treatment of Yaws-A Randomized Non Inferiority Trial in Ghana. Johnson C, editor. PLoS Negl Trop Dis. 2017; 11(1):e0005154.

23. Mitjà O, Lukehart SA, Pokowas G, Moses P, Kapa A, Godornes C, et al. Haemophilus ducreyi as a cause of skin ulcers in children from a yaws-endemic area of Papua New Guinea: a prospective cohort study. Lancet Glob Heal. 2014; 2(4):e235–41.

24. Houinei W, Godornes C, Kapa A, Knauf S, Mooring EQ, González-Beiras C, et al. Haemophilus ducreyi DNA is detectable on the skin of asymptomatic children, flies and fomites in villages of Papua New Guinea. Lammie PJ, editor. PLoS Negl Trop Dis 2017;11(5):e0004958.

25. González-Beiras C, Marks M, Chen CY, Roberts S, Mitjà O. Epidemiology of Haemophilus ducreyi Infections. Emerg Infect Dis. 2016; 22(1):1–8.

26. Grange P a., Gressier L, Dion PL, Farhi D, Benhaddou N, Gerhardt P, et al. Evaluation of a PCR test for detection of Treponema pallidum in swabs and blood. J Clin Microbiol. 2012;50(3):546–52.

27. Marks M, Chi K-H, Vahi V, Pillay A, Sokana O, Pavluck A, et al. Haemophilus ducreyi associated with skin ulcers among children, Solomon Islands. Emerg Infect Dis. 2014; 20(10):1705–7.

28. Taleo F, Macleod CK, Marks M, Sokana O, Last A, Willis R, et al. Integrated Mapping of Yaws and Trachoma in the Five Northern-Most Provinces of Vanuatu. Centurion-Lara A, editor. PLoS Negl Trop Dis. 2017;11(1):e0005267.

29. Marks M, Mitjà O, Vestergaard LS, Pillay A, Knauf S, Chen C-Y, et al. Challenges and key research questions for yaws eradication. Lancet Infect Dis. 2015; 15(10):1220–5.

